# Cryptic transmission and re-emergence of Cosmopolitan genotype of Dengue Virus Serotype 2 within Ho Chi Minh City and Southern Vietnam

**DOI:** 10.1101/2023.04.17.23288515

**Authors:** Vi T. Tran, Rhys P. D. Inward, Bernardo Gutierrez, Nguyet M Nguyen, Isabelle Rajendiran, Phong Nguyen Thanh, Tam Cao Thi, Kien T.H. Duong, Moritz U. G. Kraemer, Sophie Yacoub

## Abstract

**Background:** Dengue virus (DENV) is a major global public health threat and causes substantial morbidity and mortality in hyperendemic countries like Vietnam with its largest city, Ho Chi Minh City (HCMC), recently experiencing its largest DENV outbreak in over a decade. Despite this, there is currently a lack of data on circulating DENV serotypes/genotypes and monitoring of transmission dynamics within HCMC, which presents a challenge for the design and implementation of effective DENV mitigation strategies.

**Methods:** We generated 45 DENV envelope (E) gene sequences from human plasma samples collected in southern Vietnam between 2017 and 2022. We applied phylogenetic methods to infer the probable route of virus introductions into HCMC and its surrounding areas, as well as their approximate timing, using additional sequences from Southern and South-east Asia.

**Findings:** We found evidence of the co-circulation of multiple DENV serotypes/genotypes and the re-emergence of the DENV-2 Cosmopolitan genotype in southern Vietnam. Furthermore, we detected at least three independent seeding events of the Cosmopolitan genotype into Vietnam, the earliest of which is estimated to have occurred two years before the earliest sampling date, providing evidence of at least two seasons of cryptic transmission.

**Interpretation:** Our findings emphasize the urgent need for comprehensive DENV surveillance in HCMC and Vietnam to guide appropriate and effective public health responses and improve understanding of recent outbreak dynamics in Vietnam and neighbouring countries. Such efforts may also help predict epidemic dynamics of DENV in future seasons.

**Funding:** This work was supported by the Wellcome Trust [106680] and Wellcome Trust [226052/Z/22/Z]

**Evidence before this study:** Dengue virus (DENV) is a significant threat to global health causing high levels of morbidity and economic damage. With the limited surveillance infrastructure for DENV, particularly during the COVID-19 pandemic, little is currently known about the transmission dynamics in Ho Chi Minh City (HCMC) and Vietnam presenting a challenge for the design and implementation of DENV mitigation strategies.

**Added value of this study:** Genomic epidemiology is a powerful approach to gain insights into the spatio-temporal dynamics of viruses and to detect new viral variants. Our analyses shows the existence of multiple co-circulating DENV-2 genotypes in southern Vietnam with multiple distinct and continued introductions of the Cosmopolitan genotype into HCMC over multiple years.

**Implications of all the available evidence:** Findings from this study will assist local and regional dengue surveillance and control programs as well as adding to our understanding on DENV genomic epidemiology and transmission dynamics. Critically, current dengue screening and surveillance methodology should be modified to enable detection of these novel lineages. The emergence of these genotypes and their impact on dengue evolution need to be explored on a larger scale. These results will also allow vital missing genomic data to be incorporated into models used for importation dynamics analysis. We plan to integrate these findings into a DENV forecasting tool being developed as part of Wellcome funded multidisciplinary project - DART (Dengue Advanced Readiness Tools), which aims to build an integrated digital system for dengue outbreak prediction and monitoring.

## Introduction

Dengue virus (DENV, family Flaviviridae) is one of the most important vector-borne viruses globally with more than 2·5 billion people living in regions of high risk of infection, with the majority of those at risk coming from inter-tropical countries such as Vietnam.^1^ Globally, DENV causes an estimated 105 million cases annually, with around three-quarters of DENV infections being asymptomatic,^2^ and causes significant economic damage.^3^ Symptoms of DENV infection can range from mild, self-limited acute febrile illness to severe organ dysfunction and shock.^4^

DENV can be classified into four genetically and antigenically distinct serotypes, named DENV-1 to 4, which in turn have diversified into distinct genotypes with varying degrees of geographical persistence.^5^ DENV-1 evolved into five distinct genotypes, named I–V, while DENV-2 is divided into six genotypes some of which are named after their historical geographical regions of detection: Asian/American, Asian I, Asian II, Cosmopolitan, American^6^ and sylvatic (the latter referring to transmission cycle rather than geographical distribution, but generally found in West Africa and Southeast Asia).^7^ DENV-3 and DENV-4 are categorised into four (I-IV) genotypes each.^6^ As an arbovirus, DENV is predominantly transmitted through *Aedes aegypti* mosquito vectors, and secondarily by *Ae. Albopictus.*^8^ It features transmission cycles that take place exclusively among human populations;^9^ the reemergence of viruses circulating in non-human species can occur but is rare,^7^ and could be an influential component in some epidemic settings.^10^

Within Vietnam, DENV is common in both urban and peri-urban settings across the country with an estimated 1·6 million cases per year.^3^ In the southern provinces of Vietnam, there is persistent year-round transmission (hyperendemicity) with epidemics peaking during the rainy season between June and November.^11^ In the north of the country, in the regions surrounding Hanoi, DENV has been emerging for the past 25 years, with seasonal interruptions to local transmission during the cooler winter months and significant year-to-year fluctuations in outbreak intensity. Previous studies have shown that all four DENV serotypes have circulated at some point in Vietnam, as is common amongst hyperendemic countries, with DENV-1 and DENV-2 being the most frequently detected serotypes (Fig. 1B).^12^

**Fig. 1.**
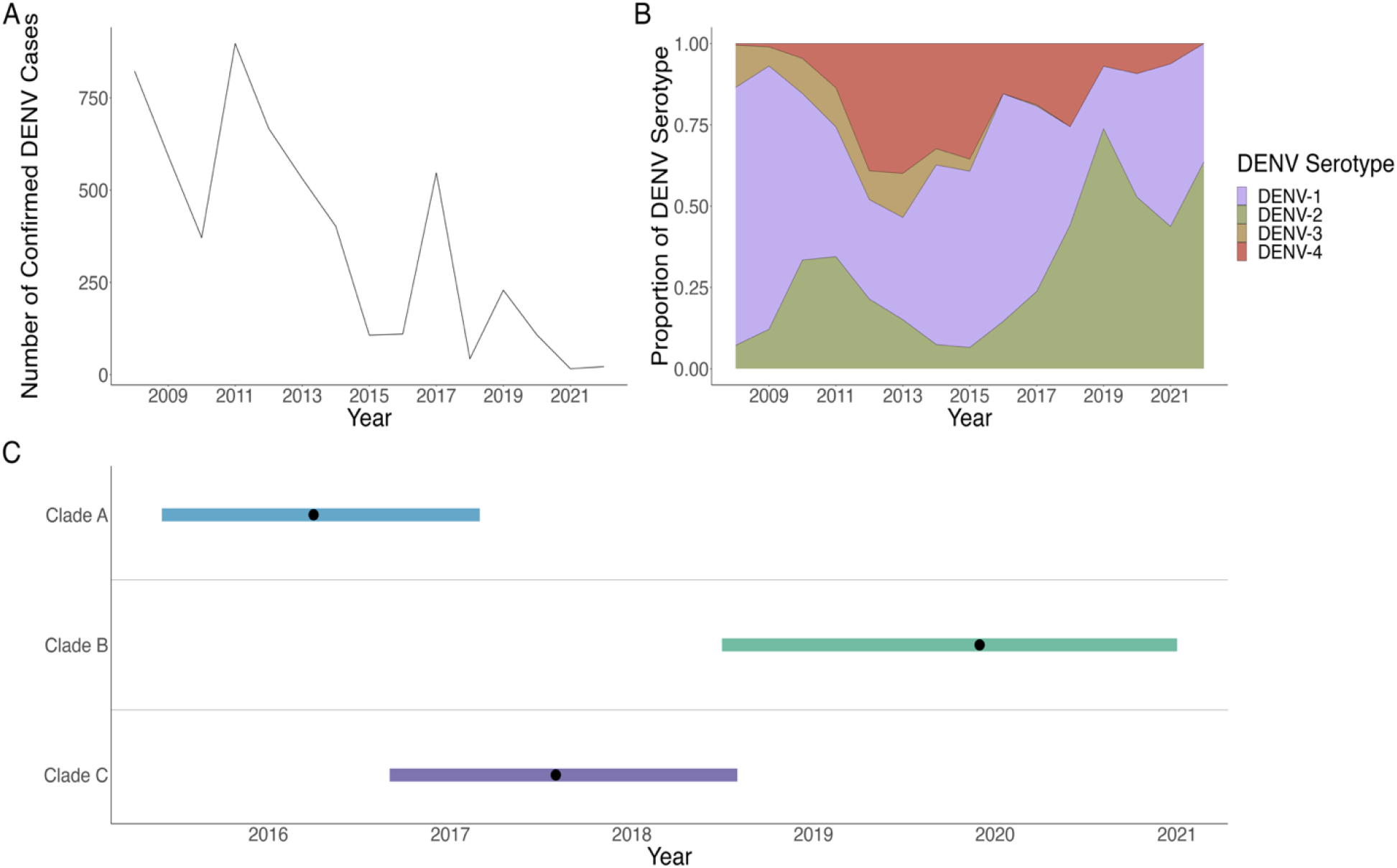
Times series of confirmed cases by NS1 enrolled within studies conducted by OURCU-HCMC (A), the proportion of cases assigned to each serotype in HCMC taken from studies conducted at OURCU-HCMC (B), and the inferred TMRCA (black dot) of the first transmission event by each lineage (Clade A, B, C) and their 90% CI (shaded area) by the Cosmopolitan genotype within Vietnam (C)

In Ho Chi Minh City (HCMC) and its surrounding provinces, DENV-1 had been the dominant serotype until 2018, when DENV-2 replaced the former as the dominant serotype (Fig. 1B). These serotype replacement dynamics are common for DENV and can be partially explained by cross-protective immunity between serotypes^13,14^ and intra-serotypic genetic diversification.^15^ Genotype replacements within a single serotype were also observed in the early 2000s in HCMC and Vietnam in general: the DENV-2 Asian I genotype replaced the previously dominant Asian/American genotype between 2003 and 2006, after which it became the predominant DENV-2 genotype.^12^ The Cosmopolitan genotype was circulating at low frequencies in HCMC during 2006 (1.07% of all DENV-2 viruses) but no further reports from Vietnam were made after 2011, despite it being prevalent in other parts of Asia, America and Africa.^16–18^

During 2022, HCMC experienced a large DENV outbreak, with 78,561 dengue confirmed cases by the HCMC Centre for Disease Control from January 1st to December 11^th^.^19^ This represents a 5-fold increase compared to the number of confirmed cases reported within the same period in 2021.^19^ Asymptomatic cases of DENV may lead to underreporting of the actual number of cases. Many factors may be associated with this large outbreak including climatic factors, reduced population immunity, and the introduction of new serotypes or genotypes, as well as the possible diversification of currently circulating lineages. There is currently a lack of data on DENV serotype/genotype dynamics within HCMC and Vietnam, which presents a challenge for the design and implementation of DENV mitigation strategies. As such, this study describes the recently circulating DENV-2 genotypes/lineages in HCMC and its surrounding areas. Since DENV accumulates a measurable number of mutations over time, even on the time scale of a few weeks, there is a sufficient resolution from genomic data to infer epidemiologically relevant characteristics of the virus, namely the transmission routes of these new DENV-2 lineages into Vietnam from bordering and other South and Southeast Asian countries and the times at which these events occurred. This will improve our understanding of the recent dengue outbreak dynamics in Vietnam and contribute not only in terms of dengue control for public health in Vietnam but also to help investigate their transmission dynamics across Vietnam and neighbouring countries.

## Methods

We produced 45 DENV-2 viral envelope (E) gene sequences from human plasma samples, originally collected from 362 laboratory-confirmed dengue patients, who participated in studies at the Hospital for Tropical Diseases in HCMC, Vietnam, between 2017 and 2022. Of these, 303 (83·7%) were positive with qRT-PCR assay for dengue, DENV-2 was predominant (72·3%), whereas DENV-1 and DENV-4 respectively accounted for 23·1% and 4·6% of positive samples. No samples were positive with DENV-3. Amongst the 219 DENV-2-positive samples, we randomly selected 45 samples with high or medium viremia levels that gave clear bands on agarose gel electrophoresis (median Cp-value for the samples was 25·53 [IQR=23·38-27·29]). The patients’ median age was 26 (IQR=17·5-33·5) years. Viral RNA from each sample was isolated using MagNA Pure 96 DNA and Viral NA Small Volume Kit (Roche, 06543588001) in the MagNA Pure 96 System.

DENV RNA was subjected to cDNA synthesis using SuperScript IV First-Strand Synthesis System (Cat # 18091200, Thermo Fisher Scientific) on a Mastercycler system (Eppendorf, Germany) according to the manufacturer’s instruction. Mix1 containing 5μL of viral RNA, 1μL of random hexamer primer and 1μL of a mixture of 10mM deoxyribonucleotide triphosphates (dNTPs). A mixture of 7μL of Mix 2 containing 4μL of 5x Reverse Transcript buffer, 1μL of each RNAse inhibitor and Superscript IV Reverse Transcriptase was added to Mix 1 and make up a final volume of 20μL. cDNA was synthesised by the thermocycler program in Supplementary Table 1.

PCR reactions to amplify the E gene were performed as 3-tube reactions using a high fidelity DNA polymerase (Cat # M0491L, New England Biolabs). The PCR mix contained 1μL dNTPs (0·4mM), 2μL of eluted primers containing both forward and reverse primers (10μM), 3μL of cDNA, and 0·5μL of DNA polymerase (2000 units/ml) diluted to a final volume of 25μL per reaction. The amplification reactions were performed according to the manufacturer’s instructions (Supplementary Table 2). PCR amplification products were visualised by agarose gel electrophoresis. The primer sets are available in Supplementary Table 3.

We purified PCR products using magnetic beads (Cat # A63882, Beckman Courter) and quantified nucleic acid concentration using a NanoDrop™ 2000/2000c Spectrophotometer (Cat # ND-2000). Sanger sequencing of purified PCR products was carried out for samples with a nucleic acid concentration greater than 10 ng/μL. Sequencing of the complete E gene was achieved by 3 overlapping amplicons. The sequencing reactions were performed with the BigDye Terminator v3.1 Cycle Sequencing Kit (Cat # 4337455) according to the manufacturer’s instructions (Supplementary Table 4). The reaction was terminated by adding 2μL of 3M sodium acetate and 100mM EDTA. A DNA precipitation step was then carried out with absolute ethanol (97%) and washed with ethanol 70%, and samples were then resuspended in 10μL of Hi-Di formamide. Sanger sequencing was performed using an Applied Biosystems 3130/3130xl Genetic Analyzer. Individual E gene consensus sequences were generated using the CLC Genomic Workbench version 9.0.

### Data collation, sequence alignment, and tree building

We collected 6013 of both DENV-2 whole genome (WG) and E gene sequences along with their associated metadata from Southern and Southeast Asia (SEA) between 2010-01-01 and 2022-12-19 from GenBank (data downloaded 2022-12-19).^20^ We removed any sequences with missing metadata, such as the absence of collection date or location, resulting in a total of 5836 sequences (which include 139 sequences from Vietnam). For sequences with incomplete metadata, i.e., yyyy or yyyy-mm, we took the mid-point of that year (e.g., 2011 → 2011-06-15). To facilitate the visualisation of phylogenies, we classified the sequence collection location information into four discrete categories: OUCRU-HCMC Vietnam, Other Vietnam, Border (countries that share a border with Vietnam I.e., China, Laos, Cambodia) and other South and SEA (countries that don’t share a border with Vietnam).

Due to errors in the serotype labelling within the GenBank metadata, we utilised the Arbovirus typing tool from Genome Detective to identify the serotype and genotype of each sequence.^21^ All entries that were not DENV-2 were removed. We aligned sequences to the reference sequence (NC_001474.2) using MAFFTV.7.^22^ All sequences were trimmed from position 937 to 2421 to ensure only the E gene was included. Additionally, sequences were further excluded if they had >5% missing or ambiguous sites, leaving a dataset of n = 4497. Lastly, we created a second dataset (n = 2812) containing sequences designated Cosmopolitan by Genome Detective.

After the initial data processing, the cleaned sequence alignment files were used to construct a maximum-likelihood (ML) tree for all DENV-2 genotypes and a separate phylogeny for the Cosmopolitan genotype alone. ML-trees were inferred using IQTREE2^23^ with a non-parametric ultrafast bootstrap to estimate node support and a Shimoda-Hasegawa approximate likelihood ratio test (SH-aLRT)^24^ to estimate branch support (command line: Iqtree2 -s -bb 1000 -alrt 1000). We used a TIM2 nucleotide substitution model with empirical base frequencies and a free rate model for rate heterogeneity with four rate categories based on a hierarchical model selection procedure as implemented in the ModelFinder application.^25^ The output ML trees were assessed for temporal signal by selecting the root position that maximises R^2^ of the regression of collection times on genetic distances to the tree root using TempEst v1.5.3,^26^ removing outliers and re-estimating trees when necessary. An outlier was defined as any tip whose genetic distance exceeds four standard deviations of the sample mean for each year. This resulted in a final dataset of 4496 sequences for all DENV-2 and 2809 sequences for the Cosmopolitan genotype. The ML tree for the Cosmopolitan genotype was then time-calibrated, informed by tip sampling dates (yyyy-mm-dd), using TreeTime.^27^

The mugration model for ancestral node reconstruction implemented within TreeTime^27^ was used to infer the location of the ancestral node state of the Cosmopolitan genotype along the phylogeny, using the country of origin of each sequence as a discrete trait. To infer the ancestral state, TreeTime employs a maximum-likelihood framework that estimates the probability of the ancestral states at each node of the phylogeny, given the observed states of its descendants. The most likely location of each node in the phylogeny is then presented as the inferred ancestral location.

Root-to-tip regression plots for the Cosmopolitan ML tree (before time calibration) are available (Supplementary Fig.1). All tree visualisations were created using the ggtree package^28^ within the programming language R.

## Results

A total of 45 DENV E gene sequences, 29 from HCMC and 16 from other southern Vietnamese regions, were generated from samples collected from previous and ongoing studies at the Hospital for Tropical Diseases in HCMC from 2017-2022 (Fig. 1A). Serotype and genotype identification of these sequences revealed the existence of multiple co-circulating DENV-2 genotypes in southern Vietnam (Fig. 1B & 2). We found that, of the total 45 sequences, 17 belonged to the Asian-I genotype, which has been established within HCMC and southern Vietnam since 2006.^12^ The remaining 28 sequences were classified as belonging to the Cosmopolitan genotype, which has only been sporadically and infrequently detected within HCMC and southern Vietnam since 2011 but is currently prevalent in other regions of South and Southeast Asia (Fig. 3).^16–18^

**Fig. 2.**
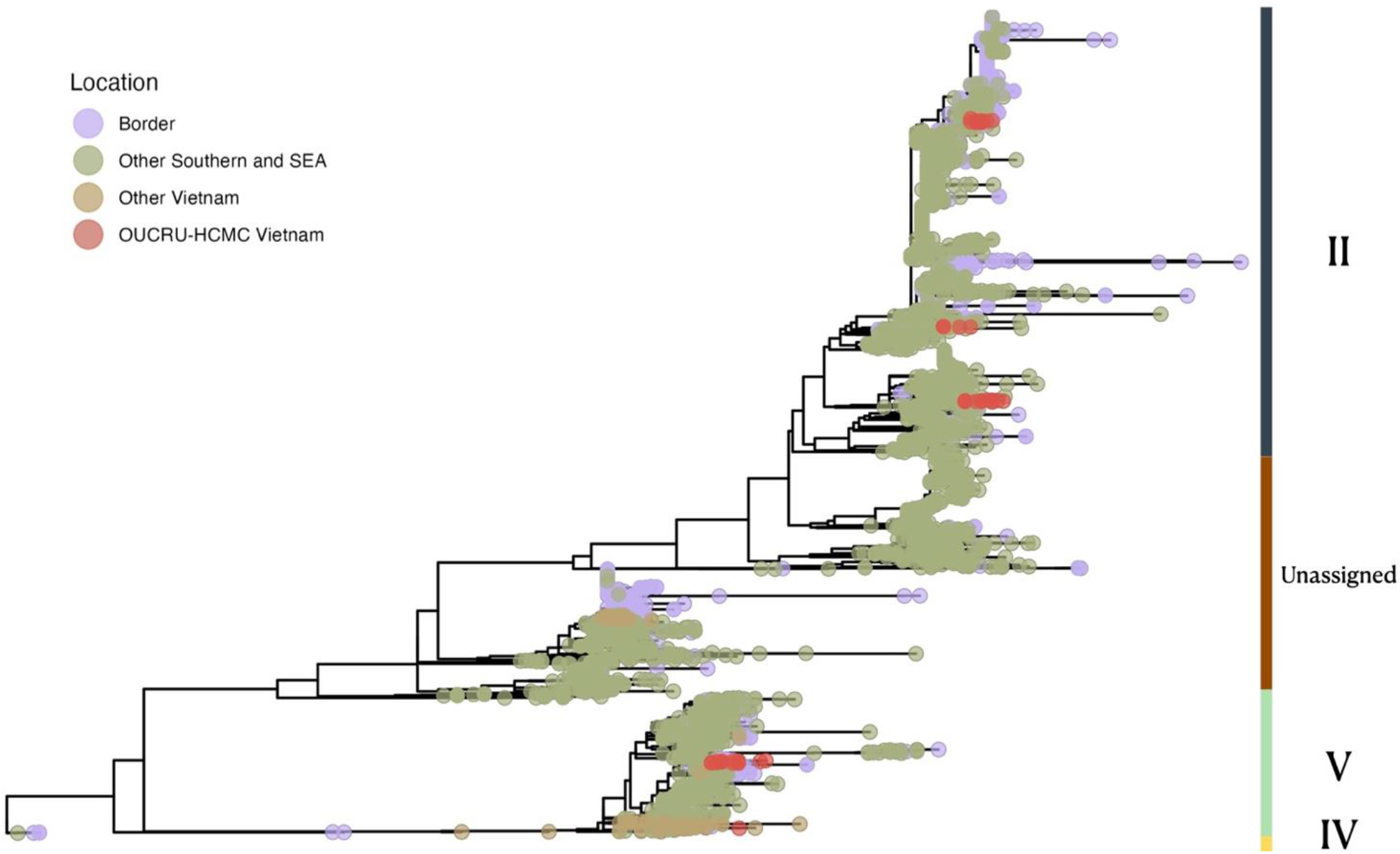
ML tree showing the genotypes of DENV-2 present within South and South-east Asia (including Vietnam) from 2010 – 2022.

**Fig. 3.**
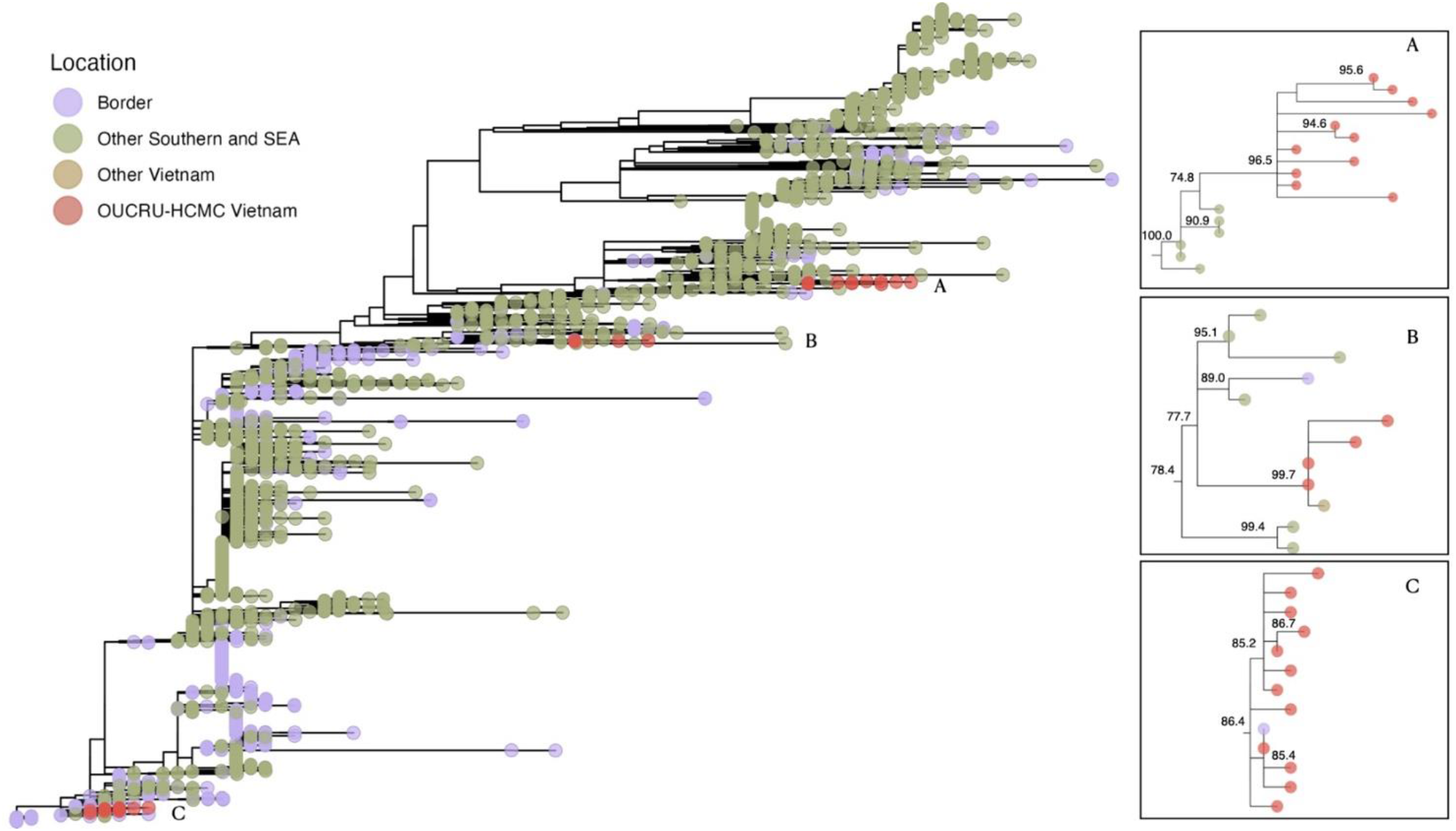
ML tree showing the Cosmopolitan genotype of DENV-2 within South and South-east Asia (including Vietnam). Boxes A-C show each transmission lineage circulating within Vietnam alongside their node support values.

While the DENV-2 genotype Asian-I has been known to circulate in southern Vietnam for over a decade,^12^ the re-emergence of the Cosmopolitan genotype represents a novel observation. An ML phylogeny of the Cosmopolitan genotype shows at least 3 distinct introduction events of the genotype into southern Vietnam (Fig. 3). In the context of this study, an introduction is defined as a monophyletic cluster of sequences (>3 sequences) from Vietnam with an inferred geographical origin outside of Vietnam according to the mugration model performed within TreeTime (i.e., the parental node inferred to have occurred in a country outside of Vietnam; Supplementary Table 5). The majority of sequences from HCMC and southern Vietnam (27/28) cluster within three distinct clades labelled A, B and C (Fig. 3). Clade A (node support 96·5%) comprises 11 sequences with an ancestral node inferred to have occurred in Indonesia (60·7% confidence). Similarly, the ancestral node of Clade B (node support 99·7%) is also inferred to have occurred in Indonesia (100·0% confidence), and it encompasses four sequences from this study alongside an additional sequence from Vietnam, of a returning traveller to China (GenBank: OP984834). In contrast, Clade C (node support 86·4%) is composed of 12 sequences, with one of these sequences from China, and its inferred ancestral node can be traced back to Cambodia (100% confidence). While individual clades show robust phylogenetic support, their internal tree topologies are poorly resolved; this is likely a consequence of incomplete sampling of transmission chains following their introduction into Vietnam. While the diversification of these clades is unclear, the sampling locations of the tips show circulation across provinces, with all clades being observed in HCMC. Clade A circulated in provinces north and east of HCMC while clades B and C were observed in provinces north and west (Fig. 4).

**Fig. 4.**
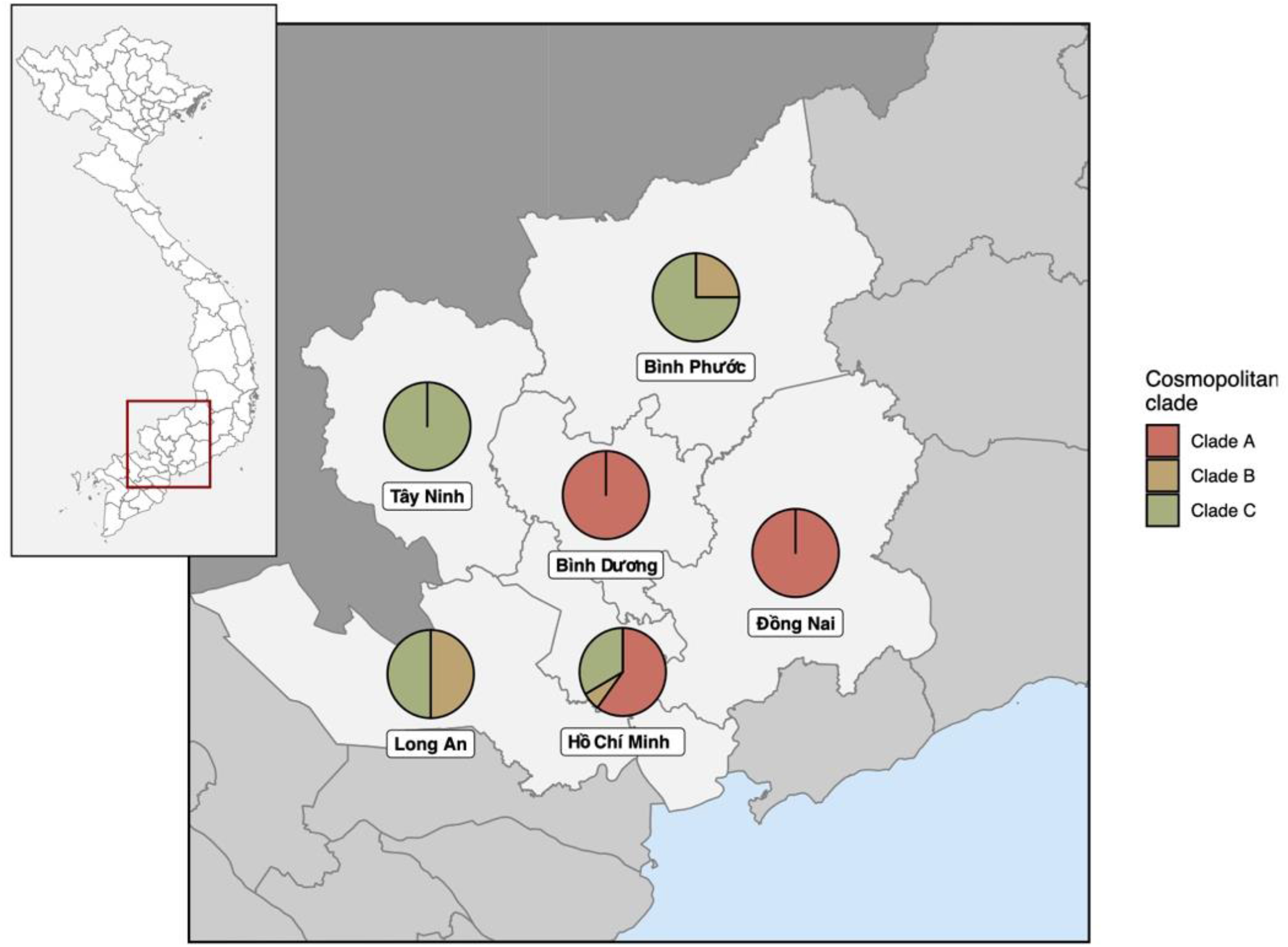
Spatial distribution of DENV-2 Cosmopolitan clades in south Vietnam. Clades A, B and C correspond to independent introductions into Vietnam from other countries and circulated amongst six provinces in south Vietnam (including Ho Chi Minh City).

Estimating the time to the most recent common ancestor (TMRCA) of the three clusters suggests that clade A (Fig. 3A) has been circulating in southern Vietnam the longest, with the clade’s earliest inferred transmission event occurring as early as April 2016 (90% confidence interval (CI): June 2015 - March 2017, Fig. 1C). This was followed by clade C (August 2017; 90% CI: August 2016 - August 2018, Fig. 1C) and clade B (December 2019; 90% CI: July 2018 - January 2021, Fig. 1C). Each of the three clusters has distinct TMRCAs suggesting that there have been continued introductions of the Cosmopolitan genotype into HCMC over multiple years (Fig. 5). Moreover, two of the clades, A and C, have shown evidence of at least half a decade of local persistence, with intervals of 1750 days (Cluster C) and 1,757 days (Cluster A) between the TMRCA and the most recent sampling date within the clade. Moreover, there were 688 days between the earliest TMRCA and the earliest sampling date amongst the three clades; this implies that there have been multiple dengue seasons of undetected cryptic transmission of the Cosmopolitan genotype within HCMC and surrounding provinces.

**Fig. 5.**
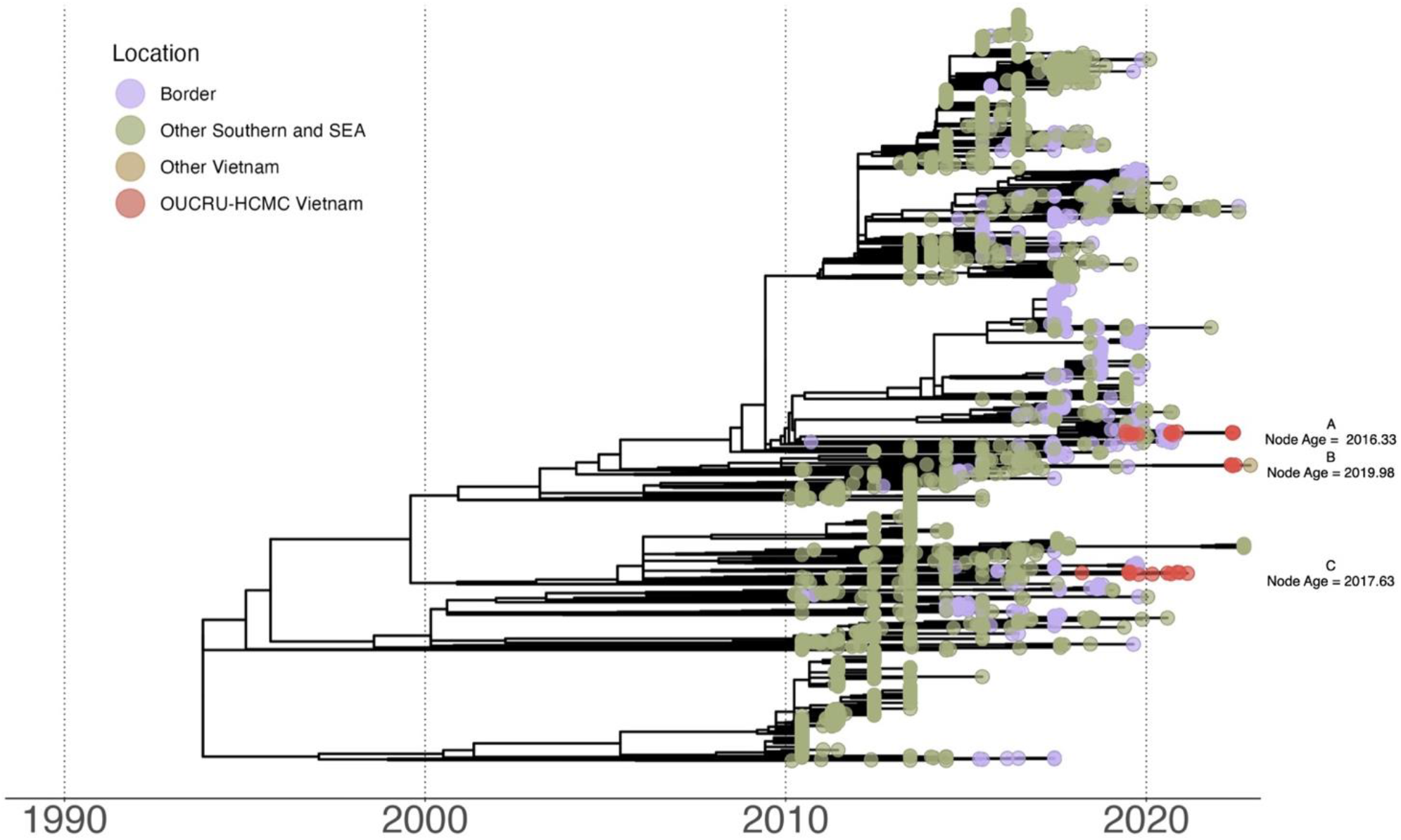
Time-scaled ML tree showing the Cosmopolitan genotype DENV-2. A-C refer to each transmission lineage and their estimated TMRCA.

## Discussion

Phylogenetic methods have been utilised in this study to analyse the available DENV-2 sequences from HCMC, enabling us to infer the probable route of introduction of the virus into the city and surrounding areas, and the approximate timing of these events.

We have found strong evidence of the co-circulation of multiple DENV-2 genotypes and the re-emergence of the Cosmopolitan genotype within HCMC. Previous studies have linked the successful emergence of a new viral lineage or serotype within a population to previous exposure and subsequent immunity to a different DENV serotype, which could create an immunological niche of individuals who are more vulnerable to the new serotype.^29^ Furthermore, the diversification of locally circulating viruses can also drive epidemic trends or the re-emergence of viral types, as has been observed during episodes of viral lineage replacement.^30^ In this context, the potential for novel importations to expand the viral genetic diversity circulating in a location can be an additional and meaningful mechanism driving DENV serotype and genotype dynamics.

Given that DENV-1 was the predominant serotype in Vietnam before 2018 (Fig. 1B), cocirculating with the DENV-2 Asian-I genotype at low frequencies, the severity and size of the current DENV outbreak in HCMC and its accompanying replacement of the dominant serotype from DENV-1 to DENV-2 could be associated with the re-emergence of the Cosmopolitan genotype.^31,32^ To this point, molecular clock analysis of the Cosmopolitan genotype within HCMC revealed multiple introductions over the last five years, at least two years before the earliest sampling date (2018-03-21) of our sequences from HCMC (Fig 1C & 3). This suggests at least two complete seasons of undetected transmission facilitated by the disruption to national surveillance programmes by the COVID-19 pandemic. The discovery of multiple introductory events (Fig. 3) indicates the current DENV dynamics within Vietnam may have been triggered by successive cycles of seeding of the Cosmopolitan genotype, thereby increasing the likelihood of establishment and sustained spread.^33,34^ Additionally, these findings raise the possibility that the Cosmopolitan genotype, which has been circulating in neighbouring countries (Fig. 2 & 3)^16–18^ but has been notably absent in Vietnam, may have been imported into the country in the past, but was unable to establish until recently. The specific mechanisms by which a genotype re-emerges following new importations and becomes established are still unclear and require further investigation.

Conducting large-scale surveillance of dengue in HCMC is challenging due to the number of asymptomatic cases^2^ and the dependence on syndromic surveillance. Also, changing climatic conditions result in highly heterogeneous dengue epidemiological trends across seasons.^35^ This poses further challenges for an effective dengue surveillance system, as successfully predicting transmission hotspots is difficult and planning resource allocation to conduct surveillance in such locations in advance becomes challenging.^36^ Moreover, limited resources for viral genomic sequencing, in addition to the diversion of these limited resources from DENV to SARS-CoV-2 surveillance during the COVID-19 pandemic, may have also exacerbated these difficulties. As a result, addressing these multiple challenges is crucial to improving DENV surveillance in HCMC.

Through our mugration analysis, we have identified both Indonesia and Cambodia as likely sources of our detected introduction events. Within both these countries, DENV is hyperendemic with year-round transmission.^37,38^ In addition, Indonesia has one of the highest dengue burdens globally.^39^ However, it is important to consider that phylogeographic inference is heavily affected by sampling biases,^40–43^ and that unsampled viral lineages can have a significant effect on the estimated sources of importations.^44^ Nonetheless, both countries are plausible sources for new imports: the availability and ease of international air travel would allow infected travellers from countries like Indonesia to arrive in well-connected regions like HCMC with established populations of *Aedes sp.* mosquitoes,^34^ while Cambodia shares a permeable border with southern Vietnam and has established transport, tourist and economic links to HCMC.^45^

This work highlights the need for and importance of continuous and systematic virus sequencing within HCMC and the surrounding provinces. The early detection of new genotypes and serotypes relies on the swift identification of novel viral lineages which can in turn be linked with ongoing DENV transmission dynamics. This is a requirement to generate timely information to inform appropriate vector control initiatives and public health responses against DENV.^46^ Overall, a comprehensive approach to surveillance and data integration is crucial for the effective management and prevention of vector-borne diseases like dengue in HCMC and can become a crucial tool in predicting the epidemic dynamics of the disease in future seasons for hyperendemic countries like Vietnam.

## Data Availability

All data used within this study are open source with details of how to obtain them found in the data availability section

## Contributors

R.P.D.I, V.T.T, S.Y, B.G.G, and M.U.G.K. developed the idea and planned the research. V.T.T collected the samples and performed sequencing. R.P.D.I, B.G, and I.R developed the bioinformatic pipeline with R.P.D.I conducting the analyses. R.P.D.I, V.T.T, S.Y, B.G, and M.U.G.K. wrote the first draft of the manuscript. K.T.H.D contributed to samples sequencing procedure and initial data cleaning. N.M.N contributed to areas related to clinical knowledge and patient recruitment. P.N.T and T.C.T contributed to management and oversight the study conduction in the hospital. All authors interpreted the data, contributed to writing, and approved the manuscript.

## Data sharing statement

All genomic data can be found here: https://www.ncbi.nlm.nih.gov/genbank/ (Genbank Acknowledgements in Supplementary Table 6). Code to reproduce the analyses presented in this study are freely available at https://github.com/rhysinward/Phylodynamics-HCMC.git.

## Acknowledgement

Role of the funding sources: R.P.D.I and M.U.G.K acknowledges funding from European Union Horizon 2020 project MOOD (#874850). M.U.G.K and B.G acknowledges acknowledge funding from the Oxford Martin School Pandemic Genomics programme.M.U.G.K. acknowledges funding from the Wellcome Trust, a Branco Weiss Fellowship, The Rockefeller Foundation and Google.org. V.T.T, K.T.H.D, N.M.N and S.Y acknowledges funding from The Wellcome Trust (106680). The contents of this publication are the sole responsibility of the authors and do not necessarily reflect the views of the European Commission or the other funders.

## Supplement Information

**Supplementary Table 1.**
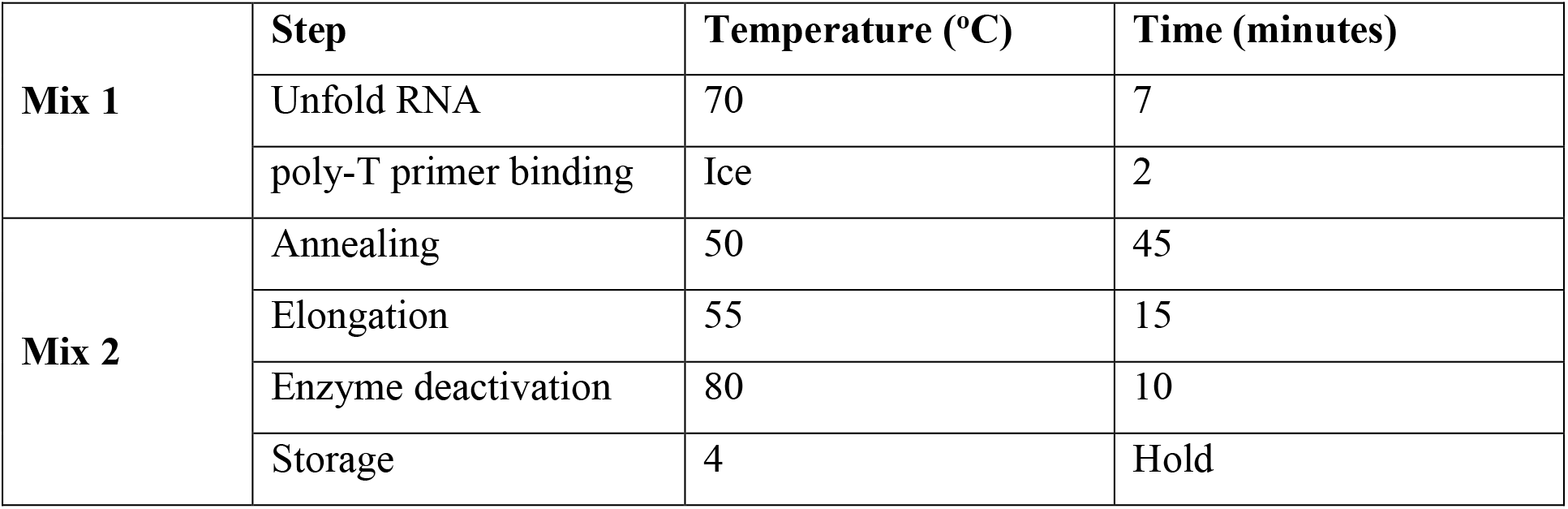
Thermocycler program for cDNA synthesis using SuperScript IV First-Strand Synthesis System (Cat # 18091200, Thermo Fisher Scientific)

**Supplementary Table 2.** Thermocycler program for PCR DENV with high fidelity DNA polymerase (Cat # M0491L, New England Biolabs)

| Step | Temperature (°C) | Time | Cycles |
| --- | --- | --- | --- |
| Initial Denaturation | 98 | 30 secs | 1 |
| Denaturation | 98 | 10 secs | x 40 |
| Annealing | 60 | 1.5 min |  |
| Extension | 72 | 30 secs |  |
| Final extension | 72 | 10 mins | 1 |
| Storage | 4 | Hold |  |

**Supplementary Table 3.**
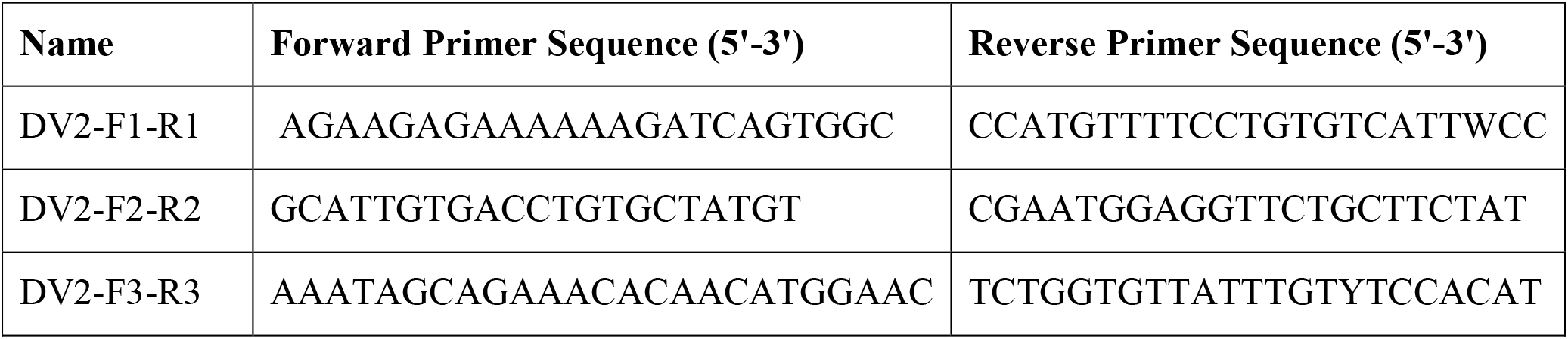

**Supplementary Table 4.** Thermocycler program for Sequencing using the BigDye Terminator v3.1 Cycle Sequencing Kit (Cat # 4337455)

| Step | Temperature (oC) | Time | Cycles |
| --- | --- | --- | --- |
| Initial Denaturation | 96 | 1 min | 1 |
| Denaturation | 96 | 10 secs | x 35 |
| Annealing | 50 | 10 secs |  |
| Extension | 60 | 1 min |  |
| Storage | 4 | Hold |  |

**Supplementary Fig. 1.**
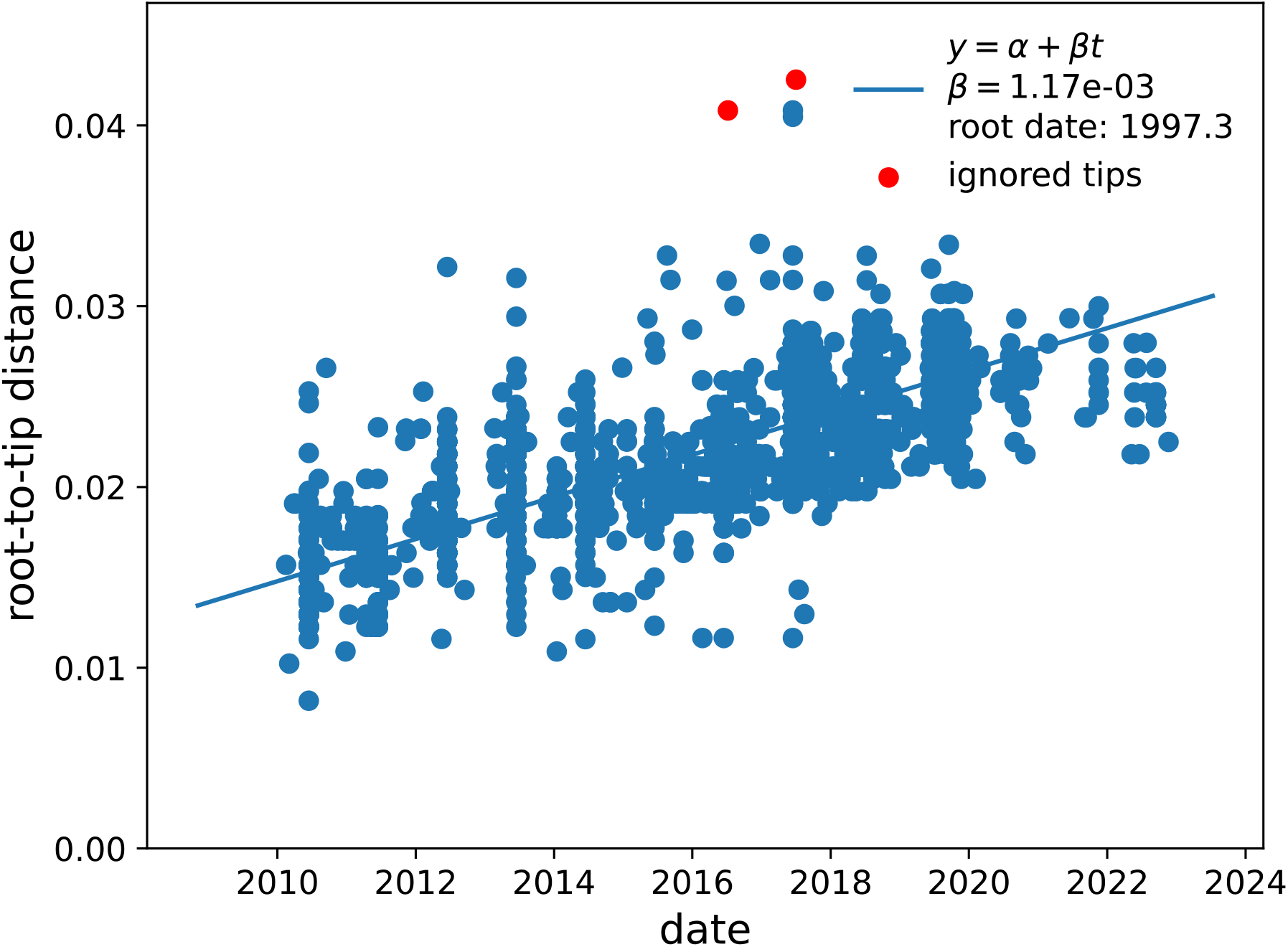
Root-to-tip regression by TreeTime of the Cosmopolitan DENV-2 dataset

**Supplementary Table 5** Results from mugration analysis can be found here - https://github.com/rhysinward/Phylodynamics-HCMC/blob/main/results/confidence.csv

**Supplementary Table 6** GenBank acknowledgement table can be found here - https://github.com/rhysinward/Phylodynamics-HCMC/blob/main/Data/sequences.csv

